# Causal associations between type 1 diabetes mellitus and cardiovascular diseases: A Mendelian randomization study

**DOI:** 10.1101/2023.07.07.23292393

**Authors:** Zirui Liu, Haocheng Wang, Zhengkai Yang, Yu Lu, Cao Zou

## Abstract

**Background:** The presence of type 1 diabetes mellitus (T1DM) has been demonstrated to pose an increased risk for developing cardiovascular diseases (CVDs). However, the causal relationships between T1DM and CVDs remain unclear due to the uncontrolled confounding factors and reverse causation bias of the observational studies.

**Aim:** To investigate the causal relationships between T1DM and seven major CVDs, including myocardial infarction (MI), heart failure (HF), coronary artery disease (CAD), atrial fibrillation (AF), coronary atherosclerosis, peripheral atherosclerosis, and stroke, using a two-sample bidirectional Mendelian randomization (MR) method.

**Method:** We selected genetic instruments for T1DM and the seven CVDs from the largest available genome-wide association studies (GWAS) of European ancestry for the MR analysis. Three complementary methods: inverse variance weighted (IVW), weighted median, and MR-Egger were used for the MR estimates. The potential pleiotropic effects were assessed by MR-Egger intercept and MR-PRESSO global test. Additionally, multivariable MR (MVMR) analysis was performed to examine whether T1DM has independent effects on CVDs with adjustment of potential confounding factors. Moreover, a two-step MR approach was used to assess the potential mediating effects of these factors on the causal effects between T1DM and CVDs.

**Results:** Causal effects of T1DM on peripheral atherosclerosis (odds ratio [OR]=1.06, 95% confidence interval [CI]: 1.02–1.10; *p* = 0.002)] and coronary atherosclerosis (OR=1.03, 95% CI: 1.01–1.05; *p* = 0.001) were found. The results were less likely to be biased by the horizontal pleiotropic effects (both p values of MR-Egger intercept and MR-PRESSO Global test > 0.05). In the following MVMR analysis, we found the causal effects of T1DM on peripheral atherosclerosis and coronary atherosclerosis remain significant after adjusting for a series of potential confounding factors. Moreover, we found that hypertension partly mediated the causal effects of T1DM on peripheral atherosclerosis (proportion of mediation effect in total effect: 11.47%, 95% CI: 3.23%–19.71%) and coronary atherosclerosis (16.84%, 95% CI: 5.35%–28.33%). We didn’t find significant causal relationships between T1DM and other CVDs, including MI, CAD, HF, AF, or stroke. For the reverse MR from CVD to T1DM, no significant causal relationships were identified.

**Conclusion:** This MR study provided evidence supporting the causal effect of T1DM on peripheral atherosclerosis and coronary atherosclerosis, with hypertension partly mediating this effect.

## 1. Introduction

Type 1 diabetes mellitus (T1DM) is a prevalent chronic autoimmune disease that affects approximately 30 million individuals, accounting for 10% of all diabetes cases^1-3^. It often occurs during childhood and adolescence, with a global prevalence of around 500,000 individuals^2^. Over the years, the incidence rate of T1DM has been increasing^4^, leading to a heavy burden on families and economies.

The pathogenesis of T1DM involves the autoimmune destruction of pancreatic β-cells, resulting in an absolute insulin deficiency and resultant hyperglycemia^5^. Persistent hyperglycemia can cause damage to both the microvascular and macrovascular systems, thereby contributing to a modest decline in overall life expectancy and a significant reduction in disability-free life expectancy.^6^

Recent observational studies have indicated that patients with T1DM are at substantially higher risk of developing cardiovascular diseases (CVD) including heart failure (HF), coronary artery disease (CAD), atrial fibrillation (AF), myocardial infarction (MI), atherosclerosis (AS), and stroke^7-12^. However, the causal relationships between T1DM and CVDs remain unclear due to uncontrolled confounding factors and reverse causation bias of the observational studies. Understanding the causal relationships between these two diseases is critical for the disease prevention and management, and thus reducing the substantial disease burden.

Mendelian randomization (MR) has been proven to be a powerful approach for clarifying causal relationships using genetic variants as instrumental variables^13^. As the genetic variants were randomly segregated during meiosis and fixed during lifetime, MR minimizes the bias due to unmeasured confounding factors and reverse causation^14^. Therefore, the MR approach is conceptually similar to a randomized controlled trial (RCT) but being more widely used and cost-effective. Previous MR studies have indicated robust causal relationships between type 2 diabetes (T2D) and CVD^15-17^. However, whether T1DM plays a causal role for the development of CVD is still unclear given the differences of risk factors, etiology, and underlying genetic factors between T1DM and T2DM^4,6^. In the present study, we performed the first bidirectional two-sample MR analysis to investigate the causal relationship between T1DM and CVDs. Moreover, we assessed the causal effects while adjusting for potential confounding factors such as hypertension, Type 2 diabetes mellitus (T2DM), smoking, C-reaction protein (CRP), interleukin-6 (IL6), high-density lipoprotein (HDL), low-density lipoprotein (LDL), triglyceride, and apolipoproteins through multivariable MR analysis (MVMR). Finally, we carried out a mediation analysis to explore whether these traits mediated the causal effects of T1D on CVDs.

## 2. Materials and methods

### 2.1. Study design

The bidirectional two-sample MR analysis was conducted to identify any potential causal association between T1DM and 7 CVDs, including MI, CAD, HF, AF, AS (peripheral and coronary atherosclerosis), and stroke **(Figure 1)**. And the approach we adopted for this MR analysis was grounded on 3 fundamental assumptions **(Figure S1)**.

**Figure 1.**
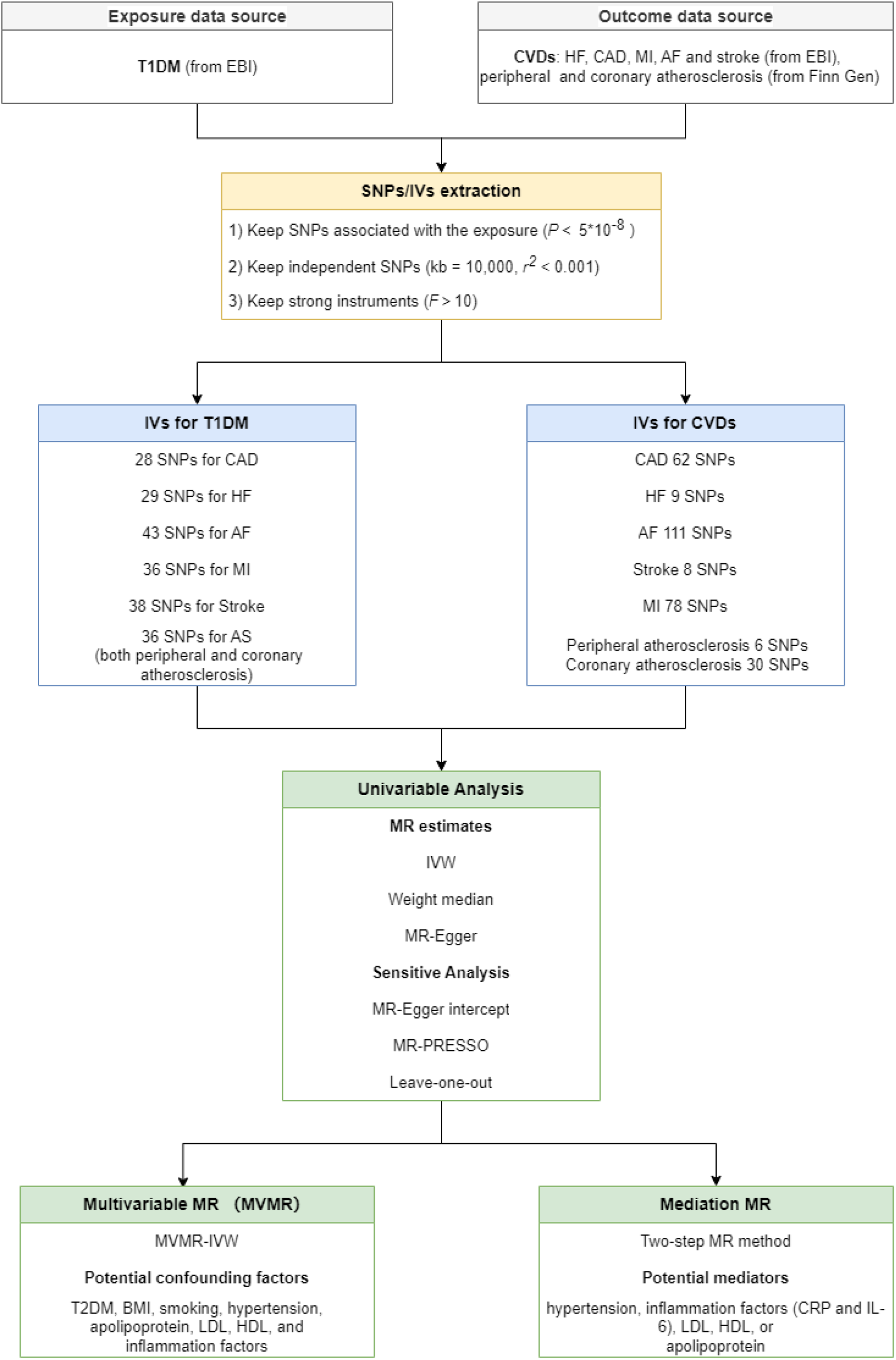
Study overview

### 2.2. Genetic association datasets

#### 2.2.1. Genetic association datasets for T1DM

Summary statistical data for T1DM with European ancestry, comprising 9,266 cases and 15,574 controls, were extracted from the European Bioinformatics Institute (EBI) database ^18^, which represents an expansive, intercontinental, and interdisciplinary data resource that remains accessible to the public within the field of life sciences^19^. To our knowledge, it is the largest scale and latest GWAS study for T1DM.

#### 2.2.2. Genetic association datasets for seven major CVDs

Our study focused primarily on investigating the causal relationship between T1DM and different types of CVDs, including MI, HF, CAD, AF, AS, and stroke. To gather the most extensive and up-to-date information on these outcomes within the European population, we selected the relevant GWAS summary studies, which are outlined in **Table 1**. The summary statistics for MI, HF, CAD, AF, and stroke were extracted from the EBI database, whereas the data pertaining to AS, including peripheral and coronary atherosclerosis, were extracted from the FinnGen Biobank, which contains genotype and phenotype data from nearly 20,000 Finnish individuals^20^.

**Table 1.**
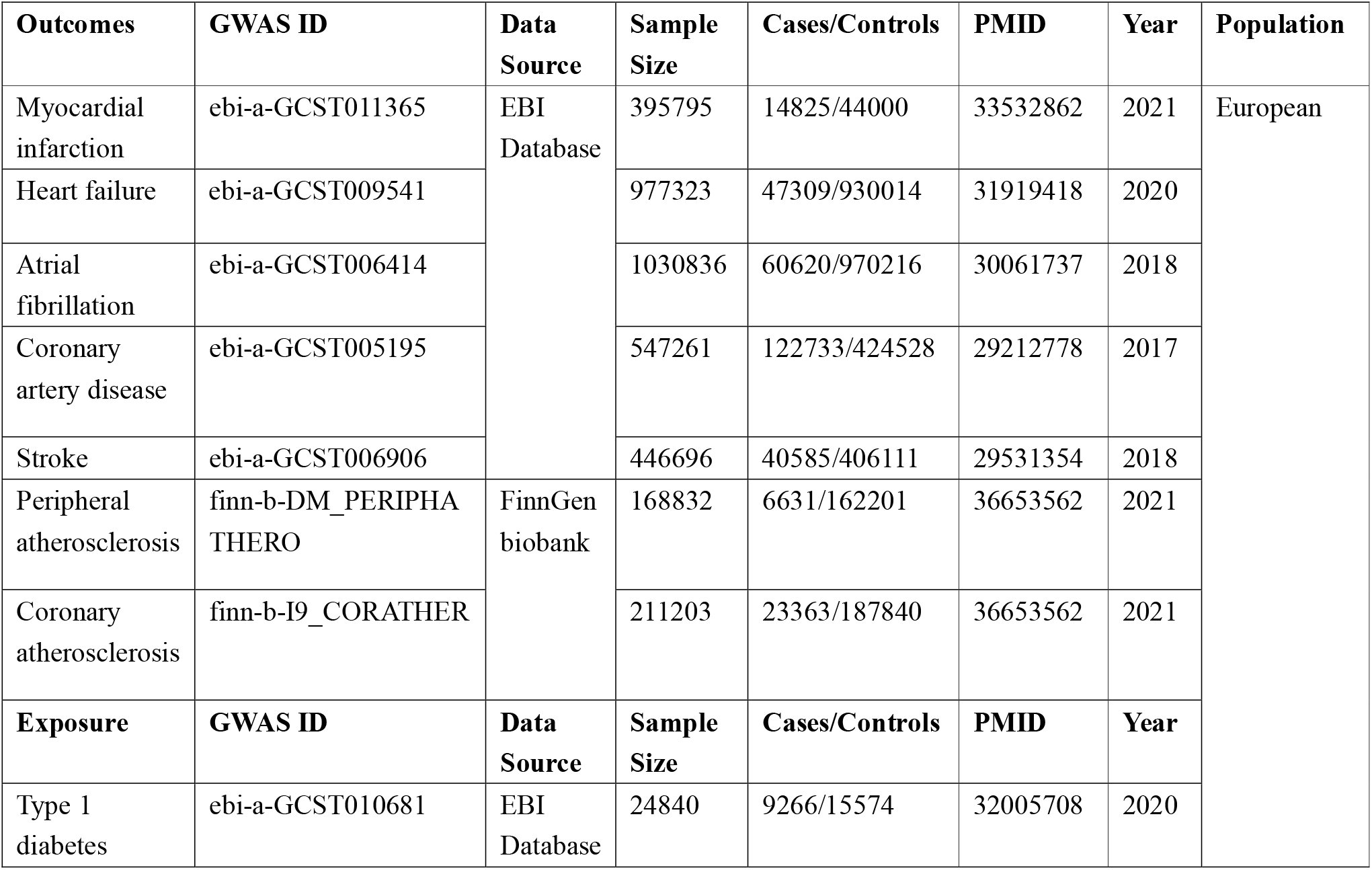
Detailed information from GWAS database.

| Outcomes | GWAS ID | Data Source | Sample Size | Cases/Controls | PMID | Year | Population |
| --- | --- | --- | --- | --- | --- | --- | --- |
| Myocardial infarction | ebi-a-GCST011365 | EBI Database | 395795 | 14825/44000 | 33532862 | 2021 | European |
| Heart failure | ebi-a-GCST009541 |  | 977323 | 47309/930014 | 31919418 | 2020 |  |
| Atrial fibrillation | ebi-a-GCST006414 |  | 1030836 | 60620/970216 | 30061737 | 2018 |  |
| Coronary artery disease | ebi-a-GCST005195 |  | 547261 | 122733/424528 | 29212778 | 2017 |  |
| Stroke | ebi-a-GCST006906 |  | 446696 | 40585/406111 | 29531354 | 2018 |  |
| Peripheral atherosclerosis | finn-b-DM_PERIPHA THERO | FinnGen biobank | 168832 | 6631/162201 | 36653562 | 2021 |  |
| Coronary atherosclerosis | finn-b-I9_CORATHER |  | 211203 | 23363/187840 | 36653562 | 2021 |  |
| Exposure | GWAS ID | Data Source | Sample Size | Cases/Controls | PMID | Year |  |
| Type 1 diabetes | ebi-a-GCST010681 | EBI Database | 24840 | 9266/15574 | 32005708 | 2020 |  |

#### 2.2.3. Informed consent statement and ethics approval statement

As the necessary consent and ethics approvals were obtained for individual studies that contributed to this MR study, no additional consent or ethics approval was required specifically for the present study.

### 2.3. IV selection

To identify the instrumental variables (IVs), we followed a two-step process. Firstly, we extracted SNPs robustly associated with the exposures (*p* < 5e-8). Secondly, we retained only the independent SNPs (kb = 10,000, *r*^*2*^ < 0.001) based on the linkage disequilibrium (LD) structure of European populations. To assess the strength of the instrumental variables, we employed the F-statistics, which is calculated as 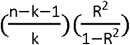, where R^2^ is the proportion of inter-individual variance explained by the instrument, n represents the sample size, and k is the number of SNPs. All the instruments used in the MR analyses were greater than the empirical threshold of 10 to minimize the potential weak instrument bias^21^.

### 2.4. Statistical analysis

We utilized the “TwoSampleMR” ^22^package for the causal estimates, and outliers were detected using the “MR-PRESSO” package. The MVMR analysis was performed using the “MVMR” and “MendelianRandomization” ^23^packages. The MR estimates were represented by odds ratios (OR) with 95% confidence intervals (CIs). All statistics were calculated using R software 4.2.2 (The R Foundation for Statistical Computing).

#### 2.4.1. Univariable MR analysis

The causal effects were estimated using the random effect inverse variance weighted (IVW) method^24^. Since the IVW method requires all the IVs to be valid in order to obtain an unbiased estimate, we also performed the MR analyses using two alternative MR methods (weighted median and MR-Egger) to assess the robustness of the results. Moreover, the potential horizontal pleiotropy was evaluated by the MR-Egger intercept and the MR-PRESSO global test.

MR-PRESSO outlier test was used to identify potential outliers. If an outlier SNP was found (p<0.05), the causal effects were re-estimated using the remaining SNPs after removing the outliers. The causal effect was considered significant if the IVW p value was less than the Bonferroni-corrected threshold (p<0.05/7=0.007) and the results from the weighted median and MR-Egger were consistent in direction.

#### 2.4.2. Multivariable MR (MVMR)

For the significant causal associations in the univariable MR analysis, the MVMR analysis was performed using the MVMR-IVW method, aiming to adjust for potential confounding factors including T2DM, BMI, smoking, hypertension, apolipoprotein, LDL, HDL, and inflammation factors.

#### 2.4.3. Mediation analysis

Given that T1DM is an autoimmune disease and previous studies have revealed that inflammatory factors and hypertension caused by T1DM might mediate the development of cardiovascular disease^5,7,12,25-28^, we performed a mediation MR analysis using the two-step MR method^29^. In the first step, we calculated the causal effect of T1DM on mediators (β_1_), and in the second step, we estimated the causal effect of mediators on CVDs (β_2_). The significance of the mediating effects (β_1*_β_2_) and the proportion of the mediation effect in the total effect were estimated using delta method.

## 3. Results

### 3.1. Univariable analysis

The MR estimated based on IVW method indicated a strong association between genetically determined T1DM and peripheral atherosclerosis (OR=1.06, 95% CI: 1.02–1.10, *p* = 0.002) as well as coronary atherosclerosis (OR=1.03, 95% CI: 1.01-1.05, *p* = 0.001) **(Figure 2)**. However, no significant associations were found between T1DM and CAD (OR=1.00, 95% CI: 0.99–1.02, *p*= 0.65), AF (OR=1.01, 95%CI: 1.00-1.02, *p* = 0.20), HF (OR=1.01, 95% CI: 0.99–1.03, *p* = 0.25), MI (OR=1.01, 95%CI: 0.99–1.02, *p* = 0.40), and stroke (OR=1.00, 95% CI: 0.98–1.01, *p* = 0.75). Two alternative MR methods: weighted median and MR-Egger show similar results **(Table 2)**. The causal relationship between T1DM and peripheral atherosclerosis and coronary atherosclerosis were less likely to be biased by the horizontal pleiotropic effects (both p values of MR-Egger intercept and MR-PRESSO Global test > 0.05) **(Table 2)**. The “Leave-one-out plot” identified that none of the SNPs dominate the estimated causal association between T1DM and CVDs (**Figure 2c-d**).

**Table 2.**
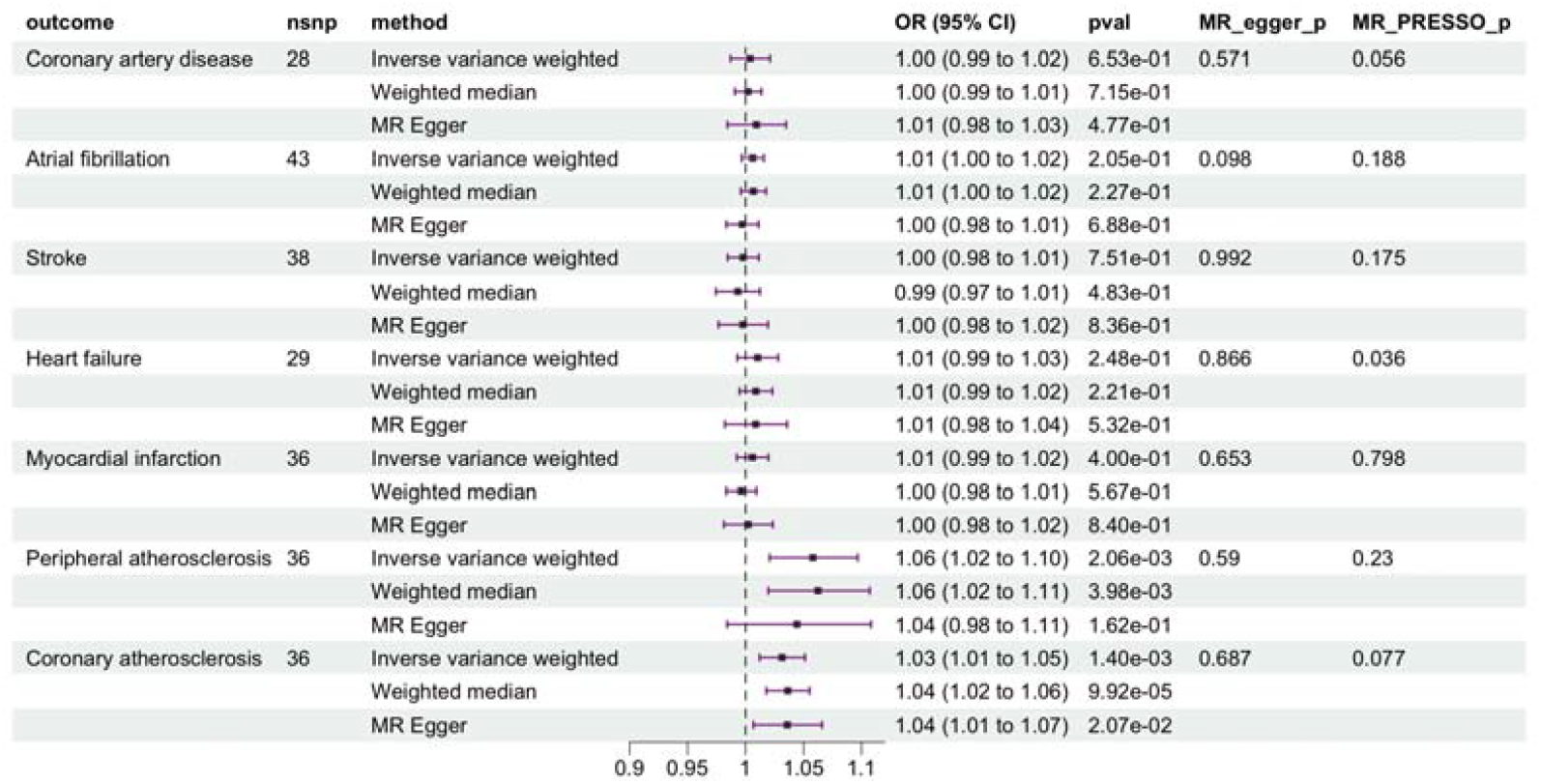
The association between T1DM and outcomes.

**Figure 2.**
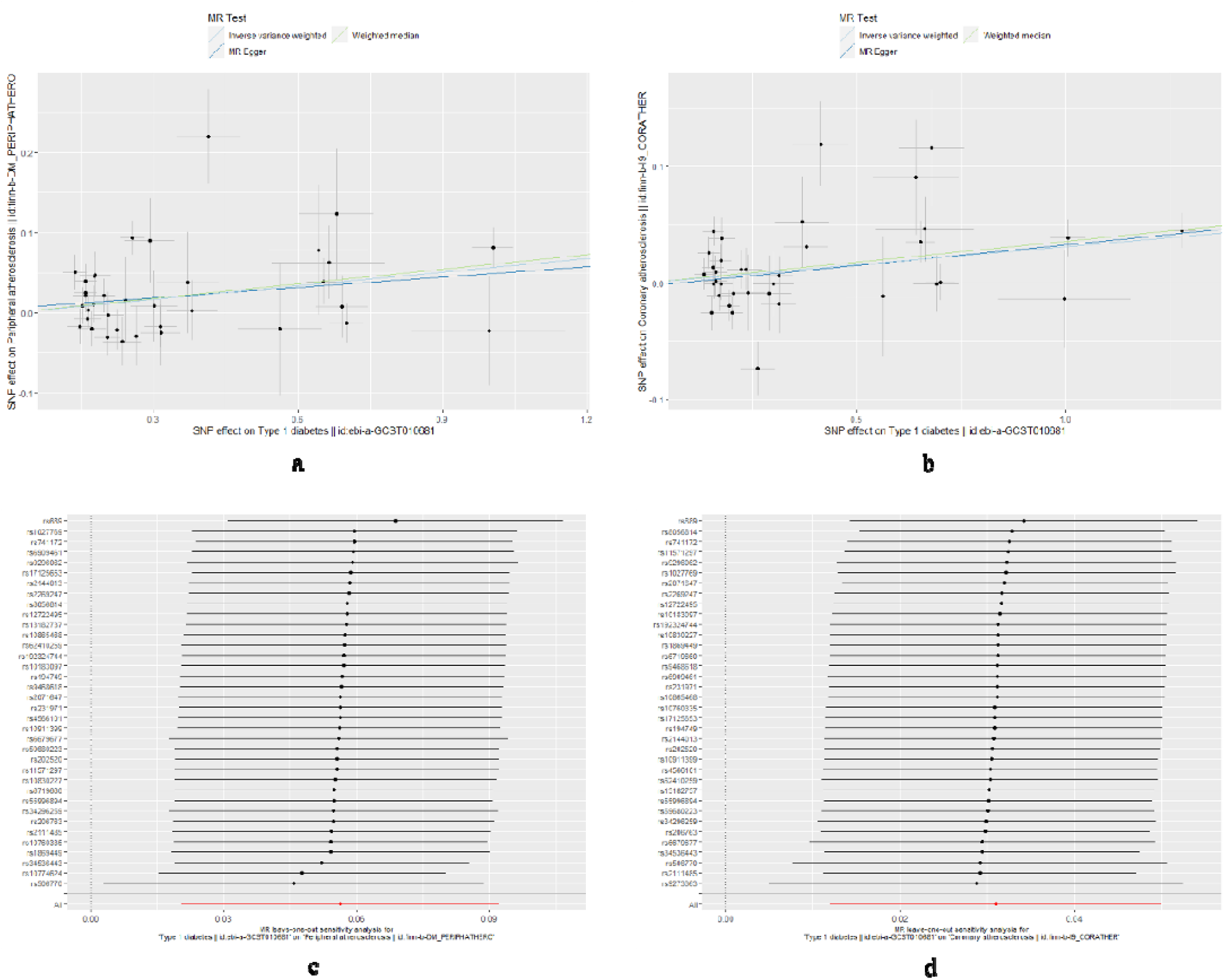
Sensitive analysis outcomes about T1DM on both peripheral and coronary atherosclerosis. Figure 2a-b showed the outcomes from three different methods. Figure 2c-d were Leave-one-out plots.

The reverse MR analyses revealed no significant causal effect of genetic predisposition to any CVDs on the risk of T1DM, including CAD (OR=1.04, 95% CI: 0.92–1.17, *p* = 0.53), AF (OR=0.98, 95% CI: 0.91-1.05, *p* = 0.59), HF (OR=0.79, 95% CI: 0.55–1.14, *p* = 0.21), MI (OR=1.07, 95% CI: 0.93–1.24, *p* = 0.35), stroke (OR=1.65, 95%CI: 0.55–4.93 *p* = 0.37), peripheral atherosclerosis (OR=2.66, 95% CI: 0.49–14.40, *p* = 0.26), and coronary atherosclerosis (OR=1.21, 95% CI: 0.81–1.80, *p* = 0.35) **(Supplementary Table S1)**.

### 3.2. MVMR analysis

The MVMR analysis was conducted to assess the direct effects of each exposure on a specific outcome. The majority of the results obtained from the univariable MR analysis were consistent with the findings from the MVMR **(Table 3)**.

**Table 3.** Multivariable MR analysis outcomes

| Potential confounding factors | Outcome | nsnp | Pval | OR | 95%CI |
| --- | --- | --- | --- | --- | --- |
| HDL | Peripheral atherosclerosis | 21 | 8.40E-07 | 1.1 | 1.06–1.15 |
|  | Coronary atherosclerosis | 21 | 0.011 | 1.04 | 1.01–1.08 |
| LDL | Peripheral atherosclerosis | 19 | 8.50E-08 | 1.13 | 1.08–1.18 |
|  | Coronary atherosclerosis | 19 | 0.002 | 1.06 | 1.02–1.09 |
| Total cholesterol | Peripheral atherosclerosis | 19 | 1.70E-07 | 1.12 | 1.07–1.16 |
|  | Coronary atherosclerosis | 19 | 0.001 | 1.06 | 1.02–1.09 |
| Triglycerides | Peripheral atherosclerosis | 21 | 8.30E-08 | 1.12 | 1.07–1.17 |
|  | Coronary atherosclerosis | 21 | 0.001 | 1.05 | 1.02–1.08 |
| apo A | Peripheral atherosclerosis | 19 | 1.60E-07 | 1.08 | 1.05–1.11 |
|  | Coronary atherosclerosis | 19 | 0.005 | 1.03 | 1.01–1.06 |
| apo B | Peripheral atherosclerosis | 25 | 3.40E-06 | 1.09 | 1.05–1.13 |
|  | Coronary atherosclerosis | 25 | 0.018 | 1.04 | 1.01–1.07 |
| Lipoprotein A | Peripheral atherosclerosis | 35 | 6.60E-06 | 1.08 | 1.05–1.12 |
|  | Coronary atherosclerosis | 35 | 0.007 | 1.04 | 1.01–1.06 |
| T2DM | Peripheral atherosclerosis | 32 | 1.70E-08 | 1.08 | 1.05–1.11 |
|  | Coronary atherosclerosis | 32 | 0.002 | 1.03 | 1.01–1.06 |
| Hypertension | Peripheral atherosclerosis | 31 | 3.00E-06 | 1.08 | 1.04–1.11 |
|  | Coronary atherosclerosis | 31 | 0.029 | 1.03 | 1.00–1.05 |
| Smoking previous | Peripheral atherosclerosis | 33 | 3.00E-06 | 1.08 | 1.05–1.12 |
|  | Coronary atherosclerosis | 33 | 0.003 | 1.04 | 1.01–1.06 |
| Smoking current | Peripheral atherosclerosis | 33 | 2.60E-05 | 1.08 | 1.04–1.12 |
|  | Coronary atherosclerosis | 33 | 0.003 | 1.04 | 1.01–1.06 |
| BMI | Peripheral atherosclerosis | 20 | 5.00E-08 | 1.1 | 1.06–1.14 |
|  | Coronary atherosclerosis | 20 | 0.001 | 1.04 | 1.02–1.07 |
| IL-6 | Peripheral atherosclerosis | 35 | 7.60E-07 | 1.09 | 1.05–1.12 |
|  | Coronary atherosclerosis | 35 | 0.001 | 1.04 | 1.02–1.06 |
| CRP | Peripheral atherosclerosis | 21 | 8.20E-09 | 1.11 | 1.07–1.15 |
|  | Coronary atherosclerosis | 21 | 0.005 | 1.05 | 1.01–1.08 |

### 3.3. Mediation MR analysis

The two-step MR was employed to perform mediation MR analysis. We aimed to investigate whether the causal relationship between T1DM and CVDs could be mediated by hypertension, inflammation factors (CRP and IL-6), LDL, HDL, or apolipoprotein. Interestingly, our findings indicated that hypertension played a role in the causal effect of T1DM on both peripheral and coronary atherosclerosis **(Table 4)**. The proportions of mediation were 11.47% (95%CI: 3.23–19.71%) and 16.84% (95%CI: 5.35–28.33%), respectively.

**Table 4.**
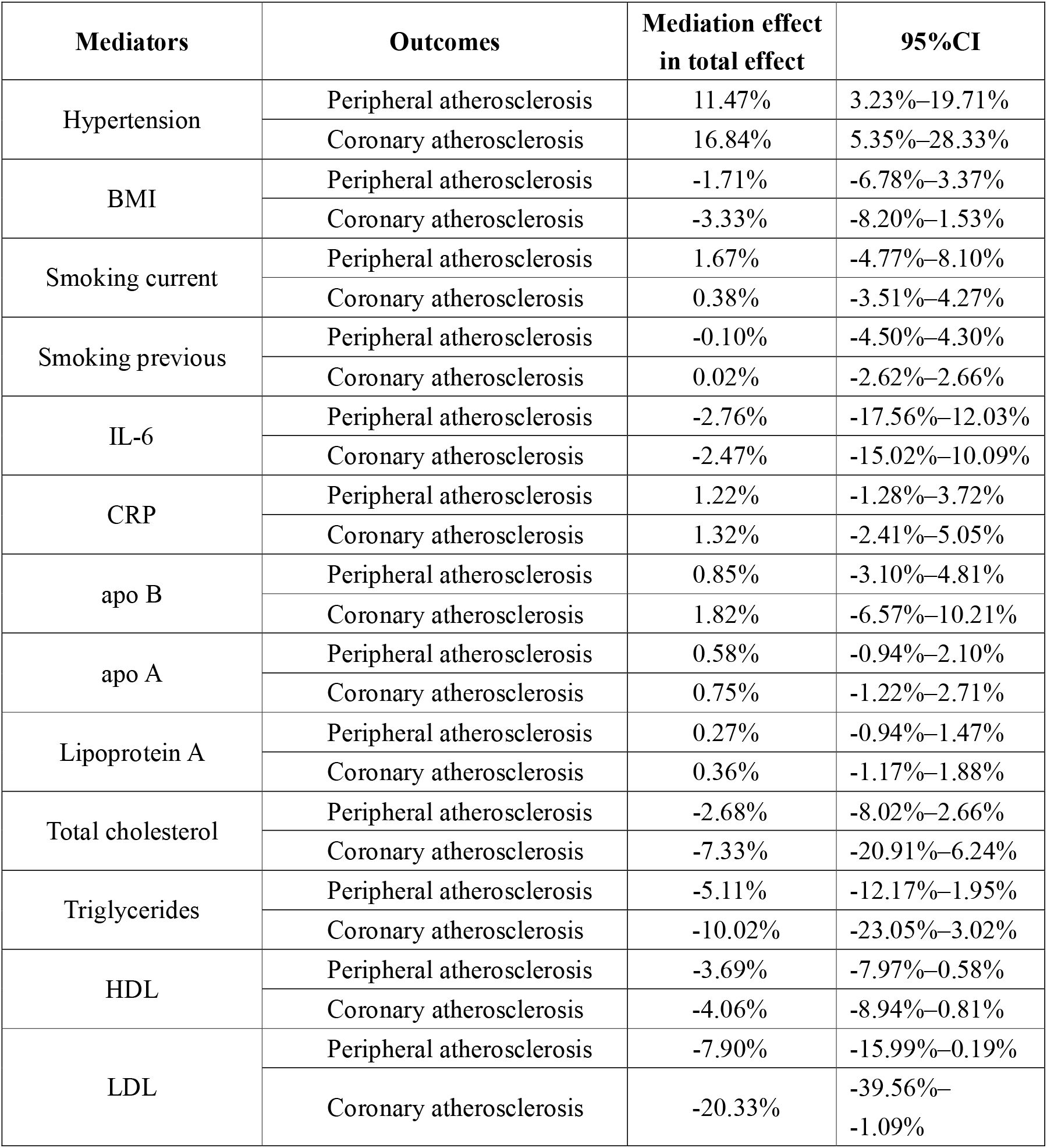
Mediation MR analysis outcomes.

## 4. Discussion

To investigate the causal relationship between T1DM in a wide range of high-frequency CVD outcomes, we conducted this MR study using large-scale GWAS summary statistics. Our analysis yielded four key findings: 1) genetic predisposition of T1DM was associated with a high risk of both peripheral and coronary atherosclerosis; 2) the causal effect of T1DM on atherosclerosis is independent of T2DM; 3) hypertension plays an important mediating role in the causal pathway from T1DM to AS; 4) No causal association was observed between T1DM and other CVDs, including HF, AF, CAD, MI, and stroke. Additionally, all the positive outcomes were validated through sensitive analysis (IVW, MR-Egger, MR-PRESSO, leave-one-out analysis, and MVMR).

The process of atherosclerosis always begins at an early stage of life among T1DM patients^8^, with endothelial dysfunction being identified as a significant pathophysiological mechanism.

Early atherosclerotic changes can be measured by both flow-mediated dilatation (FMD) and carotid intimal medial thickness(cIMT)^30,31^. A recent meta-analysis focusing on arterial damage in T1DM demonstrated a correlation between high cIMT levels and subclinical arterial injury (mean difference [d] = 0.03, 95% CI= 0.02-0.04)^32^. Mikko J. Järvisalo et.al found impaired FMD was a common manifestation in adolescents with T1DM (4.4±3.4% versus 8.7±3.6%, P<0.001)^30^.

Furthermore, inflammation factors like CRP and IL-6, as well as endothelial markers including sICAM and sVCAM, along with longitudinal lipids, may be associated with higher cIMT and lower FMD, exacerbating endothelial dysfunction and ultimately leading to atherosclerosis^30,31,33^. Recent research has identified IL-6 antagonists, such as tocilizumab and ziltivekimab, as potential therapeutic options to improve endothelial function, which can be used as preventive medication for atherosclerosis^34,35^. Therefore, antagonistic drugs targeting these molecules may potentially attenuate the progression of atherosclerosis in individuals with T1DM. However, further research is needed to explore the efficacy of antagonistic drugs targeting these molecules.

Our findings also highlighted hypertension as a major risk factor for cardiovascular complications among patients with T1DM and its role in promoting the development of atherosclerosis. The potential mechanism underlying hypertension in patients with T1DM may be attributed to diabetic nephropathy^36^. Chronic and persistent hyperglycemia, hyperlipidemia, and glomerular hypertension contribute to the deterioration of renal function^37^. Consequently, there is an accumulation of salts in the body, triggering an excessive activation of the sympathetic nervous system and renin-angiotensin-aldosterone system (RAAS), ultimately leading to hypertension^38^. This process involves various intricate molecules such as transforming growth factor (TGF β1), angiotensin 2(ANG2), vascular endothelial growth factor (VEGF), as well as signal pathways (e.g., TGFβ1-RhoA/Rho signaling)^37^. In clinical practice, inhibition of the RAAS and the use of sodium-glucose cotransporter 2 inhibitors (SGLT2i) have been validated as effective measures to protect kidney from metabolic and hemodynamic damage, which in turn helps to control blood pressure. However, further research focusing on specific cellular pathways from T1DM to diabetic kidney disease is needed and is an area of considerable interest.

In this two-sample bidirectional MR analyses, we did not find any casual relationships between T1DM and HF, MI, CAD, AF, or stroke, which is different from some previous observational studies^9-11,39,40^. A nationwide, register-based cohort study^39^ reported that T1DM patients were more susceptible to suffering from acute myocardial infarction [HR=5.77, 95% CI (4.08–8.16)], stroke[HR=3.22, 95%CI (2.35–4.42)], as well as heart failure[HR=5.07, 95% CI (3.55–7.22)]. Maryam Saeed et al. found a nine-fold excess risk of AMI in people with T1DM, HR=9.05, 95% CI (7.18–11.41))^9^. Other cohort studies have reported similar outcomes. This discrepancy can be explained as follows: 1) The outcomes drawn from observational studies are inherently affected by confounding factors. Hence, the impact of T1DM on CVDs may not be as remarkable as previously suggested. 2) Different observational studies have yielded inconsistent conclusions. For example, a population-based prospective cohort study in Sweden did not reveal a significant correlation between T1DM and AF, [HR=0.99, 95% CI 0.65–1.50)]^10^, while another study by Bin Lee Y et al. reported an opposite conclusion[HR= 1.75, 95% CI (1.53-1.99)]^11^. To sum up, the results of this MR study suggest that associations between T1DM and CVDs including HF, MI, CAD, AF, and stroke, previously reported in observational studies, may be influenced by biases such as reverse causality or confounding factors.

Extensive research has recently focused on the causal relationship between T2DM and CVD, consistently demonstrating that T2DM is a significant contributor to the development of CVD^15-17^. In our MR study, we aimed to investigate the causal relationship between T1DM and 7 high frequency CVDs, thereby expanding the knowledge in the field of diabetes and cardiovascular disease research. We specifically emphasized the causal effect of T1DM on atherosclerosis is independent of T2DM. However, our study findings did not identify any significant casual association between T1DM and HF, AF, CAD, MI, or stroke. Some potential mechanisms may partly explain the differences between T1DM and T2DM in the development of CVDs. Although chronic hyperglycemia is a common clinical manifestation and the failure of β cells is a primary event in the development of diabetes mellitus, the nature history, pathophysiologic and genetics mechanisms are all different^41^. Firstly, T1DM typically occurs in adolescence and persists throughout the lifespan, with approximately 80% of pancreatic beta cell function lost by the time of diagnosis. Conversely, T2DM is often diagnosed in middle age when patients still have more than 50% of pancreatic beta cell function^42^. Secondly, autoimmune-mediated β-cell failure leads to absolute deficiency of insulin in T1DM individuals. T cells play a critical role in inducing senescence and apoptosis of pancreatic islet β-cells^5^. On the other hand, there is no convincing evidence supporting autoimmune response in T2DM^43^. T2DM is predominantly mediated by metabolic factors, leading to a sustained decline in β-cell function and insulin resistance in the end^15^. Thirdly, recent research has focused on genetic variants associated with both T1DM and T2DM impacting β-cell^44^. The GWAS have identified more than 400 distinct genetic signals that are evidently associated with T2DM and over 50 signals influencing T1DM^45,46^, highlighting genetic differences between the two types of diabetes. In conclusion, different types of diabetes play distinct roles in cardiovascular complications in spite of the common feature of β-cells failure. More research remains to be done to develop individualized prevention strategies.

The implications of our study for clinical practice are suggested below. Firstly, as T1DM is a lifelong disease, it is significant for physicians to early and regularly evaluate atherosclerotic changes in the arterial wall, especially among adolescents. Secondly, more reasonable strategies for managing hypertension and hyperglycemia need to be developed, as they might slow down the progression of MI, CAD, AF, HF, as well as stroke. Thirdly, in order to ascertain their effectiveness, it is necessary to conduct large-scale randomized controlled trials to validate the potential of novel therapies that aim to protect β-cells in T1DM, including Imatinib^47^ and TUDCA^48^.

To the best of our knowledge, this is the first MR analysis which aims to find a causal relationship of T1DM on CVDs using the latest and largest GWAS data. We utilized the MR technique to mitigate any potential confounding bias and obtain reliable causal inference. Moreover, we employed various crucial methods to systematically investigate the presence of pleiotropy in IVs, which allowed us to address the issue of pleiotropy and enhance the reliability of the MR analysis.

However, there are still some limitations that need to be emphasized. Firstly, the majority of statistics in the GWAS were derived from individuals of European ancestry, raising concerns about the generalizability of our findings to other populations. Secondly, despite our efforts to minimize pleiotropy, it is unlikely to completely eliminate all instances of pleiotropy in Mendelian randomization studies. There may still be unrecognized pathways and confounding factors between the exposure and outcome variables, potentially introducing biases into our results. Thirdly, a potential limitation of this research is the inability to stratify the analysis based on the severity of T1DM and other important variables such as gender and age.

## 5. Conclusion

In conclusion, our MR analysis provides evidence of a causal effect of T1DM on AS, which can be mediated by hypertension. However, no convincing evidence of a causal link was found in this study between T1DM and other CVDs, including MI, CAD, HF, AF, or stroke.

## Data Availability

All data produced are available online at https://gwas.mrcieu.ac.uk/

## 8. Supplementary materials

Table S1 Causal associations of CVDs on T1DM.

Figure S1 Study design and 3 assumptions of MR analysis.

## References

1. DiMeglio LA, Evans-Molina C, Oram RA. Type 1 diabetes. Lancet. 2018;391(10138):2449–2462.

2. Katsarou A, Gudbjörnsdottir S, Rawshani A, et al. Type 1 diabetes mellitus. Nature reviews Disease primers. 2017;3:17016.

3. Green A, Hede SM, Patterson CC, et al. Type 1 diabetes in 2017: global estimates of incident and prevalent cases in children and adults. Diabetologia. 2021;64(12):2741–2750.

4. Lawrence JM, Divers J, Isom S, et al. Trends in Prevalence of Type 1 and Type 2 Diabetes in Children and Adolescents in the US, 2001-2017. Jama. 2021;326(8):717–727.

5. Vallianou NG, Stratigou T, Geladari E, Tessier CM, Mantzoros CS, Dalamaga M. Diabetes type 1: Can it be treated as an autoimmune disorder? Rev Endocr Metab Disord. 2021;22(4):859–876.

6. Cosentino F, Grant PJ, Aboyans V, et al. 2019 ESC Guidelines on diabetes, pre-diabetes, and cardiovascular diseases developed in collaboration with the EASD. Eur Heart J. 2020;41(2):255–323.

7. Sousa GR, Pober D, Galderisi A, et al. Glycemic Control, Cardiac Autoimmunity, and Long-Term Risk of Cardiovascular Disease in Type 1 Diabetes Mellitus. Circulation. 2019;139(6):730–743.

8. Harjutsalo V, Pongrac Barlovic D, Groop PH. Long-term population-based trends in the incidence of cardiovascular disease in individuals with type 1 diabetes from Finland: a retrospective, nationwide, cohort study. Lancet Diabetes Endocrinol. 2021;9(9):575–585.

9. Saeed M, Stene LC, Ariansen I, et al. Nine-fold higher risk of acute myocardial infarction in subjects with type 1 diabetes compared to controls in Norway 1973-2017. Cardiovasc Diabetol. 2022;21(1):59.

10. Larsson SC, Wallin A, Hakansson N, Stackelberg O, Back M, Wolk A. Type 1 and type 2 diabetes mellitus and incidence of seven cardiovascular diseases. Int J Cardiol. 2018;262:66–70.

11. Lee YB, Han K, Kim B, et al. Risk of early mortality and cardiovascular disease in type 1 diabetes: a comparison with type 2 diabetes, a nationwide study. Cardiovasc Diabetol. 2019;18(1):157.

12. Morandi A, Piona C, Corradi M, et al. Risk factors for pre-clinical atherosclerosis in adolescents with type 1 diabetes. Diabetes Res Clin Pract. 2023;198:110618.

13. O’Donnell CJ, Sabatine MS. Opportunities and Challenges in Mendelian Randomization Studies to Guide Trial Design. JAMA cardiology. 2018;3(10):967.

14. Sanderson E, Glymour MM, Holmes MV, et al. Mendelian randomization. Nature Reviews Methods Primers. 2022;2(1):6.

15. Huang M, Laina-Nicaise LD, Zha L, Tang T, Cheng X. Causal Association of Type 2 Diabetes Mellitus and Glycemic Traits With Cardiovascular Diseases and Lipid Traits: A Mendelian Randomization Study. Front Endocrinol (Lausanne). 2022;13:840579.

16. Liu B, Mason AM, Sun L, Di Angelantonio E, Gill D, Burgess S. Genetically Predicted Type 2 Diabetes Mellitus Liability, Glycated Hemoglobin and Cardiovascular Diseases: A Wide-Angled Mendelian Randomization Study. Genes (Basel). 2021;12(10).

17. Ahmad OS, Morris JA, Mujammami M, et al. A Mendelian randomization study of the effect of type-2 diabetes on coronary heart disease. Nat Commun. 2015;6:7060.

18. Forgetta V, Manousaki D, Istomine R, et al. Rare Genetic Variants of Large Effect Influence Risk of Type 1 Diabetes. Diabetes. 2020;69(4):784–795.

19. Rodriguez-Tomé P, Stoehr PJ, Cameron GN, Flores TP. The European Bioinformatics Institute (EBI) databases. Nucleic acids research. 1996;24(1):6–12.

20. Locke AE, Steinberg KM, Chiang CWK, et al. Exome sequencing of Finnish isolates enhances rare-variant association power. Nature. 2019;572(7769):323-328. 21.

21. Douglas Staiger JHS. Instrumental variables regression with weak instruments. econometrica. 1997;65(3):557–586.

22. Hemani G, Zheng J, Elsworth B, et al. The MR-Base platform supports systematic causal inference across the human phenome. eLife. 2018;7.

23. Yavorska OO, Burgess S. MendelianRandomization: an R package for performing Mendelian randomization analyses using summarized data. International journal of epidemiology. 2017;46(6):1734–1739.

24. Burgess S, Butterworth A, Thompson SG. Mendelian randomization analysis with multiple genetic variants using summarized data. Genetic epidemiology. 2013;37(7):658–665.

25. Pastore I, Bolla AM, Montefusco L, et al. The Impact of Diabetes Mellitus on Cardiovascular Risk Onset in Children and Adolescents. Int J Mol Sci. 2020;21(14).

26. Petrie JR, Sattar N. Excess Cardiovascular Risk in Type 1 Diabetes Mellitus. Circulation. 2019;139(6):744–747.

27. Sousa GR, Kosiborod M, Bluemke DA, Lipes MA. Cardiac Autoimmunity Is Associated With Subclinical Myocardial Dysfunction in Patients With Type 1 Diabetes Mellitus. Circulation. 2020;141(13):1107–1109.

28. Stitziel NO, Kanter JE, Bornfeldt KE. Emerging Targets for Cardiovascular Disease Prevention in Diabetes. Trends Mol Med. 2020;26(8):744–757.

29. Sanderson E. Multivariable Mendelian Randomization and Mediation. Cold Spring Harbor perspectives in medicine. 2021;11(2).

30. Jarvisalo MJ, Raitakari M, Toikka JO, et al. Endothelial dysfunction and increased arterial intima-media thickness in children with type 1 diabetes. Circulation. 2004;109(14):1750–1755.

31. Bharathy PS, Delhikumar CG, Rajappa M, Sahoo J, Anantharaj A. Early Markers of Atherosclerosis in Children and Adolescents with Type 1 Diabetes Mellitus. Indian J Pediatr. 2023;90(3):227–232.

32. Giannopoulou EZ, Doundoulakis I, Antza C, et al. Subclinical arterial damage in children and adolescents with type 1 diabetes: A systematic review and meta-analysis. Pediatr Diabetes. 2019;20(6):668–677.

33. Perez-Segura P, de Dios O, Herrero L, et al. Children with type 1 diabetes have elevated high-sensitivity C-reactive protein compared with a control group. BMJ Open Diabetes Res Care. 2020;8(1).

34. Hartman J, Frishman WH. Inflammation and atherosclerosis: a review of the role of interleukin-6 in the development of atherosclerosis and the potential for targeted drug therapy. Cardiology in review. 2014;22(3):147–151.

35. Christoffersen M, Tybjaerg-Hansen A. Targeting IL-6 in patients at high cardiovascular risk. Lancet. 2021;397(10289):2025–2027.

36. Jia G, Sowers JR. Hypertension in Diabetes: An Update of Basic Mechanisms and Clinical Disease. Hypertension. 2021;78(5):1197–1205.

37. Ricciardi CA, Gnudi L. Kidney disease in diabetes: From mechanisms to clinical presentation and treatment strategies. Metabolism. 2021;124:154890.

38. Lithovius R, Harjutsalo V, Mutter S, et al. Resistant Hypertension and Risk of Adverse Events in Individuals With Type 1 Diabetes: A Nationwide Prospective Study. Diabetes Care. 2020;43(8):1885–1892.

39. Rawshani A, Sattar N, Franzen S, et al. Excess mortality and cardiovascular disease in young adults with type 1 diabetes in relation to age at onset: a nationwide, register-based cohort study. Lancet. 2018;392(10146):477–486.

40. Kerola AM, Semb AG, Juonala M, Palomaki A, Rautava P, Kyto V. Long-term cardiovascular prognosis of patients with type 1 diabetes after myocardial infarction. Cardiovasc Diabetol. 2022;21(1):177.

41. Cnop M, Welsh N, Jonas JC, Jörns A, Lenzen S, Eizirik DL. Mechanisms of pancreatic beta-cell death in type 1 and type 2 diabetes: many differences, few similarities. Diabetes. 2005;54 Suppl 2:S97–107.

42. Srikanta S, Ganda OP, Jackson RA, et al. Type I diabetes mellitus in monozygotic twins: chronic progressive beta cell dysfunction. Annals of internal medicine. 1983;99(3):320–326.

43. Velloso LA, Eizirik DL, Cnop M. Type 2 diabetes mellitus--an autoimmune disease? Nat Rev Endocrinol. 2013;9(12):750–755.

44. Eizirik DL, Pasquali L, Cnop M. Pancreatic beta-cells in type 1 and type 2 diabetes mellitus: different pathways to failure. Nat Rev Endocrinol. 2020;16(7):349–362.

45. Onengut-Gumuscu S, Chen WM, Burren O, et al. Fine mapping of type 1 diabetes susceptibility loci and evidence for colocalization of causal variants with lymphoid gene enhancers. Nature genetics. 2015;47(4):381–386.

46. Mahajan A, Taliun D, Thurner M, et al. Fine-mapping type 2 diabetes loci to single-variant resolution using high-density imputation and islet-specific epigenome maps. Nature genetics. 2018;50(11):1505–1513.

47. Gitelman SE, Bundy BN, Ferrannini E, et al. Imatinib therapy for patients with recent-onset type 1 diabetes: a multicentre, randomised, double-blind, placebo-controlled, phase 2 trial. Lancet Diabetes Endocrinol. 2021;9(8):502–514.

48. Brozzi F, Nardelli TR, Lopes M, et al. Cytokines induce endoplasmic reticulum stress in human, rat and mouse beta cells via different mechanisms. Diabetologia. 2015;58(10):2307–2316.

