## Supplementary material for "Causal associations between type 1 diabetes mellitus and cardiovascular diseases: A Mendelian randomization study": Table S1

***Supplementary Materials***

Table S1 Causal associations of CVDs on T1DM.

| **Exposure** | **Method** | **nsnp** | **Pval** | **OR** | **95%CI** |
| --- | --- | --- | --- | --- | --- |
| Coronary artery disease | Inverse variance weighted | 62 | 0.53 | 1.04 | 0.92-1.17 |
|  | Weighted median |  | 0.66 | 0.97 | 0.83-1.13 |
|  | MR Egger |  | 0.38 | 1.11 | 0.88-1.41 |
| Atrial fibrillation | Inverse variance weighted | 111 | 0.59 | 0.98 | 0.91-1.05 |
|  | Weighted median |  | 0.25 | 0.93 | 0.83-1.05 |
|  | MR Egger |  | 0.32 | 1.08 | 0.93-1.24 |
| Stroke | Inverse variance weighted | 8 | 0.37 | 1.65 | 0.55-4.93 |
|  | Weighted median |  | 0.21 | 0.76 | 0.49-1.17 |
|  | MR Egger |  | 0.84 | 0.40 | 0-1657.37 |
| Heart failure | Inverse variance weighted | 9 | 0.21 | 0.79 | 0.55-1.14 |
|  | Weighted median |  | 0.11 | 0.72 | 0.48-1.07 |
|  | MR Egger |  | 0.48 | 0.65 | 0.22-1.98 |
| Myocardial infarction | Inverse variance weighted | 78 | 0.35 | 1.07 | 0.93-1.24 |
|  | Weighted median |  | 0.69 | 0.97 | 0.84-1.12 |
|  | MR Egger |  | 0.44 | 1.13 | 0.83-1.55 |
| Peripheral atherosclerosis | Inverse variance weighted | 6 | 0.26 | 2.66 | 0.49-14.40 |
|  | Weighted median |  | 0.97 | 1.00 | 0.83-1.21 |
|  | MR Egger |  | 0.62 | 0.13 | 0-251.40 |
| Coronary atherosclerosis | Inverse variance weighted | 30 | 0.35 | 1.21 | 0.81-1.80 |
|  | Weighted median |  | 0.81 | 0.98 | 0.86-1.12 |
|  | MR Egger |  | 0.60 | 0.78 | 0.31-1.95 |

Table S1 Causal associations of CVDs on T1DM.
