## Supplementary material for "Causal associations between type 1 diabetes mellitus and cardiovascular diseases: A Mendelian randomization study": Figure S1

***Supplementary Materials***

Figure S1 Study design and 3 assumptions of MR analysis.

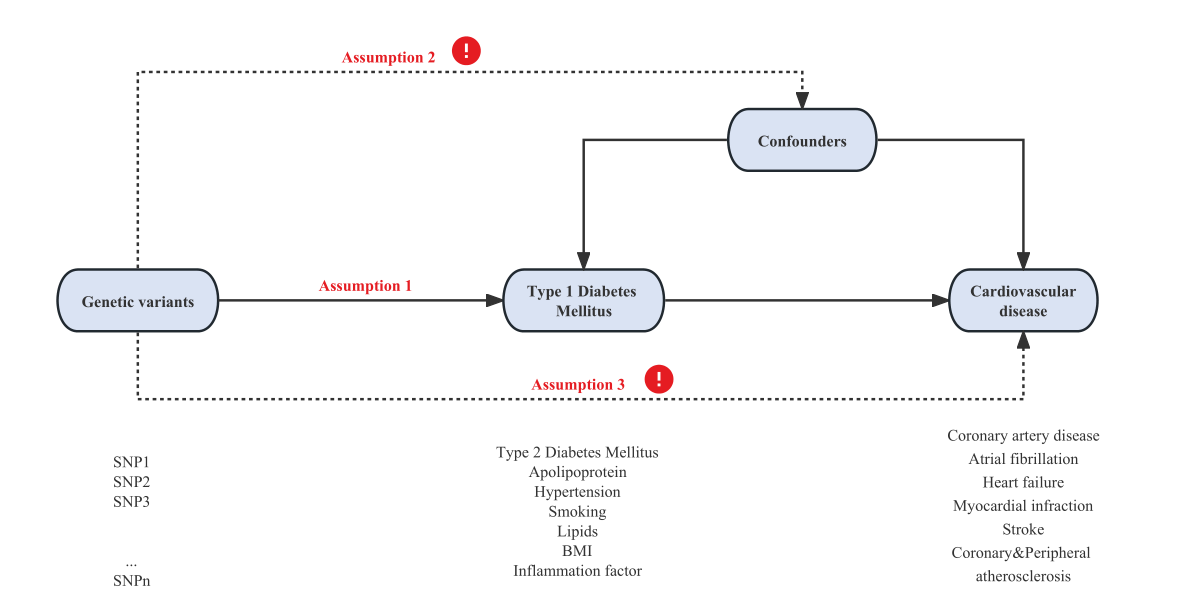

Figure S1 Study design and 3 assumptions of MR analysis.
